# PROTOCOL: LLM-Generated CONSORT Report for Increased Reporting: A Parallel-Arm Randomized Controlled Trial

**DOI:** 10.64898/2026.04.15.26350926

**Authors:** Karl Rohe, Auden Nordberg Krauska, Gary Collins, Sara Schroter

**Affiliations:** University of Wisconsin-Madison; University of Birmingham; British Medical Journal

## Abstract

**Background:** Randomized controlled trials (RCTs) often have incomplete methods reporting despite widespread adoption of the CONSORT guideline. The editorial process is supposed to detect these shortcomings and request clarifications from authors, which is time-consuming. We developed an LLM-based CONSORT Rohe Nordberg Report that highlights which CONSORT items appear fully or partially reported and checks page references claimed by authors, and then creates follow up questions for authors to more easily correct missing information.

**Methods:** This parallel-arm, superiority RCT will randomize eligible RCT submissions (after desk screening) 1:1 into intervention (editorial team and authors receive the Rohe Nordberg Report) or control (standard editorial review only). The primary outcome is whether manuscripts improve their reporting of CONSORT items in the Methods and Results sections between the original submission and first revision. This will be assessed by blinded human reviewers who evaluate the textual changes for improvements between the original and revised manuscripts for each relevant CONSORT item. Secondary outcomes include time to editorial decisions, rejection and non-resubmission rates, if authors can correctly identify where CONSORT items are reported, and extent of revisions. Human evaluators will be blinded to whether the manuscript was in the intervention or control group.

**Discussion:** By providing authors and the editorial team with specific follow up questions for each underreported CONSORT item, we hypothesize that basic underreporting will be more efficiently detected and corrected. Using blinded human reviewers as the primary outcome assessors ensures a rigorous, unbiased evaluation. If successful, this approach may help align manuscripts more closely with CONSORT standards, ultimately benefiting evidence synthesis.

**Trial Registration:** *[To be registered prior to enrollment; e.g., ClinicalTrials.gov or ISRCTN]*

**Administrative Information:** *Title:* LLM-Generated CONSORT Rohe Nordberg Report for Increased Reporting: Protocol for a Parallel-Arm Randomized Controlled Trial. IRB registered name: LLM-Generated CONSORT Report Phase III Trial

*Trial Registration:* This trial will be registered before enrollment commences in a publicly accessible registry (e.g., ClinicalTrials.gov or ISRCTN). The trial identifier will be inserted here upon registration. All items from the World Health Organization Trial Registration Data Set will be provided at the time of registration, including: primary registry and trial ID, date of registration, secondary IDs, source of funding, contact for public and scientific queries, title, research ethics review, study design, study setting, interventions, eligibility criteria, primary outcome, key secondary outcomes, target sample size, recruitment status, and results dissemination plan.

*Protocol Version:* Version 1.0 26 March 2026 *[Subsequent amendments will be tracked by version number, date, and a summary of changes.]*

*Funding:* Open Philanthropy, grant title “From Manual to Machine: Validating and Scaling LLM-Based CONSORT Compliance Assessment for Evidence-Based Medicine Publishing” Participating journals provide in-kind editorial resources (staff time, system access) to facilitate trial conduct.

*Roles and Responsibilities:* Protocol contributors
Auden Nordberg Krauska (University of Wisconsin-Madison): conceived the study design, wrote the initial protocol draft
Karl Rohe (University of Wisconsin-Madison): primary investigator, co-developed the LLM-based CONSORT and RoB 2 systems
Gary Collins (University of Birmingham): senior methodologist, contributed to trial design and statistical analysis plan
Sara Schroter (British Medical Journal): research editor, contributed to trial design and implementation plans
Hyunseung Kang (University of Wisconsin-Madison): aided in statistical analysis plan Trial sponsor
University of Wisconsin-Madison, Department of Statistics, 1300 University Avenue, Madison, WI 53706. Role of sponsor and funders
The research team leads trial design, data collection, analysis, interpretation, and reporting. The sponsor and funders have no role in data collection, management, analysis, interpretation of data, writing of the report, or the decision to submit the report for publication. The funder provides financial support only. Participating journal editorial teams are consulted for feasibility and operational feedback but do not have authority over data analysis or reporting. Trial oversight groups
The trial will be coordinated by Karl Rohe and Auden Nordberg Krauska (University of Wisconsin-Madison), with methodological input from Gary Collins (University of Birmingham). Day-to-day operations, including manuscript tracking, diff file preparation, and data management will be carried out by Auden Nordberg Krauska and trained undergraduate research assistants at UW-Madison. Sara Schroter, Research Editor at The BMJ, will facilitate integration with the journal’s editorial workflow and monitor recruitment progress. This group will meet as needed to review trial progress and resolve operational issues.

## 2. Introduction

### 2.1 Background and Rationale

Incomplete methodological details in randomized controlled trials hinder replication and can undermine subsequent evidence synthesis efforts. The CONSORT (Consolidated Standards of Reporting Trials) checklist (Hopewell et al., 2025) provides a standardized way for authors to present crucial methodological information, but adherence remains inconsistent and checking for omissions is burdensome.

Associate editors (AEs) often rely on their own scrutiny, peer reviewers, or specialized checklists to identify omissions. However, time pressures and variable expertise can lead to overlooked issues. Using a large language model (LLM) to generate a concise report of missing CONSORT items and corresponding follow up questions could make it easier for AEs to request clarifications from authors before publication.

Previous studies have evaluated the impact of CONSORT endorsement by journals on reporting quality, generally finding modest improvements (Blanco, 2020). However, no randomized trial has evaluated whether providing editors with automated, item-level CONSORT feedback with follow up questions at the manuscript level improves reporting completeness at first revision.

### 2.2 Explanation for Choice of Comparators

Standard editorial review (without any LLM-based tool) is the most direct comparator for determining whether an LLM-based editorial aid confers additional benefit. This conventional approach is well understood by editors and authors, ensuring that any observed differences can be primarily attributed to the LLM’s structured feedback rather than other sources of variation.

### 2.3 Objectives

#### Primary objective

To evaluate whether providing the editorial team with an automated Rohe Nordberg Report (vs. standard editorial approach alone) increases the proportion of first-revision manuscripts that fully report all relevant CONSORT items.

#### Secondary objectives

- Compare editorial outcomes (rejection rates, time to final decisions, proportion of manuscripts never resubmitted) between arms.
- Assess how often page-number references in authors’ submitted CONSORT checklists are correct, quantifying the system’s ability to validate these references automatically.
- Measure extent of revisions (word count of differential changes between original submission and first revision) as a proxy for authors’ efforts to address missing CONSORT items.

### 2.4 Trial Design

This is a parallel, two-arm, superiority randomized controlled trial with a 1:1 allocation ratio. Eligible RCT manuscripts that pass initial editorial screening (i.e., are not desk-rejected) at participating journals will be allocated to either:

- **Intervention arm:** The AE receives a Rohe Nordberg Report generated by the LLM-based system that mimics a peer review report. The AE may integrate that feedback into revision requests for authors at their discretion.
- **Control arm:** Standard editorial process only, with no LLM-based report provided to the AE.

After authors submit the first revision, a packet of three documents is generated for each arm and provided to blinded human reviewers: (1) the original submission, (2) the first revision, and (3) a diff file comparing the two. When both documents are submitted in Microsoft Word format, the diff file is generated using Word’s built-in Compare function (Review → Compare). When either document is in a non-Word format, both are first converted to plain text and the diff file is generated using the Unix diff utility. Reviewers assess whether the manuscript improved its reporting for each relevant CONSORT item in the Methods and Results sections. The framework is superiority: we hypothesize that the intervention arm will show greater improvement in CONSORT reporting at first revision.

## 3. Methods: Participants, Interventions, and Outcomes

### 3.1 Study Setting

The trial will be conducted within the editorial systems of The BMJ and BMJ Open [EDIT TO ADD ANY ADDITIONAL JOURNALS]. All manuscript processing occurs through the journals’ online submission and peer-review platforms. The study therefore has an international scope, encompassing RCT manuscripts submitted from any country.

### 3.2 Eligibility Criteria

#### Inclusion criteria

- Manuscripts describing a completed RCT, as identified by study screening by an LLM on the manuscript’s title and abstract.
- Submitted during the trial recruitment window to a participating journal.
- Manuscripts that pass initial desk screening (i.e., not desk-rejected).

#### Exclusion criteria

- Non-RCT study designs or manuscripts mislabeled as RCTs.
- Trials requiring rapid or expedited review that precludes the additional feedback step (e.g., urgent public health concerns).
- Resubmissions of manuscripts previously randomized in this trial.

Note: Because the unit of randomization is the manuscript (not a human participant), eligibility criteria for individual participants do not apply in the traditional sense.

### 3.3 Interventions

#### Intervention arm: LLM-Generated Rohe Nordberg Report

Upon assignment to the intervention arm, the LLM-based system processes the submitted manuscript and generates a Rohe Nordberg Report structured as a peer-review-style document. The report is designed to be actionable for both AEs and authors and contains the following elements:

- **(a) Overview:** A summary of how many of the 42 CONSORT checklist items are fully reported, how many have gaps, and how many are not applicable to the study design.
- **(b) Prioritized follow-up questions:** For each item identified as underreported, the report generates a specific, manuscript-contextualized follow-up question. Questions are organized into three tiers based on their significance for reporting transparency: Key Gaps (highest priority; cannot evaluate without), Notable Gaps (meaningful additional detail; replication is impaired), and Clarifications (minor completeness issue).
- **(c) Prompt for the author to address the underreporting:** each question cites the relevant passage from the manuscript, explains why the information matters for readers, and includes a concrete suggestion for how and where in the manuscript the gap could be addressed (e.g., "Please add 2–3 sentences to the Methods section describing…")

The report is generated from three or more independent LLM evaluations of the manuscript, which are synthesized into a consensus assessment. The follow-up questions are then refined for specificity and tone. The AE may forward the report to authors as part of the revision request, incorporate selected questions into a response to the author, and/or use it as a reference when drafting editorial feedback. The report is advisory; the editorial team retains full editorial authority over how (and whether) to act on its contents.

#### Control arm — Usual Practice

The editorial team and peer reviewers rely on their standard processes and the authors’ self-reported CONSORT checklists, without an automated LLM summary of incomplete items or page-number verifications.

##### Criteria for discontinuing or modifying allocated interventions

Not applicable. The intervention is a single report delivered to the AE at one time point and cannot be modified after delivery. Manuscripts withdrawn by authors prior to first revision, or manuscripts from journals that withdraw from the trial, will be documented and handled as described in Section 4 (Study Populations) of the Statistical Analysis Plan.

#### Strategies to improve adherence to intervention protocols

Adherence will be monitored by recording whether the AE toggled the “share with author” option in ScholarOne and whether the decision letter referenced any LLM-generated questions using the exact same wording.

#### Relevant concomitant care and interventions

Standard peer review processes will proceed in parallel for both arms. Editors and AEs cross arms and may use any other editorial aids or checklists as part of their normal workflow.

### 3.4 Outcomes

#### Primary outcome

Improvement in CONSORT reporting of Methods and Results items between the original submission and first revision, as assessed by blinded human reviewers.

For each manuscript, a diff file will be generated that displays the textual changes (insertions, deletions, and modifications) between the original submission and the first revised manuscript. Blinded human reviewers will examine these diff files and, for each CONSORT item that falls within the Methods or Results sections, will evaluate whether the manuscript increased, maintained, or reduced its reporting of that item. The specific CONSORT items evaluated will include items 9-27.

The primary analysis metric is the mean number of CONSORT items per manuscript for which human reviewers judge that reporting improved at first revision, compared between the intervention and control arms. This differs from prior studies on CONSORT compliance (Turner, 2012), as it does not place an emphasis on defining what constitutes “full reporting” versus “partial reporting” vs “incomplete reporting.” Since CONSORT was written as a guidance to authors and not as an evaluative tool, different studies on CONSORT compliance evaluate based on different criteria for the same items, making it difficult to compare compliance studies. By focusing on improvement on each item, it also makes it easier for human evaluators to code and reduces the effect of editors rejecting manuscripts that have poor reporting being a driver for the treatment effect.

##### Blinding of outcome assessment

The diff files provided to reviewers do not contain any information that could reveal arm assignment. Reviewers will not know whether the manuscript’s AE received the Rohe Nordberg Report. Reviewer allocation to manuscripts will be managed centrally to avoid systematic biases.

##### Reviewer training and reliability

Human reviewers will receive standardized training using a detailed coding manual with exemplar diff files and worked examples. Each diff file will be evaluated independently by at least two reviewers. Inter-rater reliability will be assessed using Cohen’s kappa on all double-coded manuscripts. Disagreements will be resolved by consensus discussion or adjudication by a third reviewer.

#### Secondary outcomes

1. **Correct page references:** The proportion of CONSORT items for which the authors’ stated page numbers are confirmed correct by the LLM-based system, aggregated by arm.
2. **Editorial decisions:** Rejection rate after peer review; proportion of manuscripts never resubmitted after initial decision; time (in days) from submission to final editorial decision.
3. **Author resubmission time:** Time between decision given to authors and first revision resubmission; time (in days).
4. **Extent of revisions:** Word count of differential changes (words deleted plus words added) between the original submission and first revision, as a proxy for authors’ efforts to include missing CONSORT items.
5. **Inter-rater reliability:** Agreement statistics (Cohen’s kappa) among human reviewers assessing the diff files, reported overall and per CONSORT item.

### 3.5 Participant Timeline

The schedule of enrollment, intervention delivery, and outcome assessment is as follows:

| Step | Activity | Intervention Arm | Control Arm |
| --- | --- | --- | --- |
| 1 | Submission & desk screening | Eligible manuscript identified | Eligible manuscript identified |
| 2 | Randomization (1:1) | Allocated to intervention | Allocated to control |
| 3 | Intervention delivery | AE receives Rohe Nordberg Report | Standard editorial process |
| 4 | Peer review & decision | Standard + LLM-informed feedback | Standard process only |
| 5 | First revision received | Authors revise manuscript | Authors revise manuscript |
| 6 | Diff generation | Structured diff file created | Structured diff file created |
| 7 | Blinded human review | Reviewers assess CONSORT improvement | Reviewers assess CONSORT improvement |
| 8 | Outcome determination | Data recorded | Data recorded |

*Figure: Participant (manuscript) timeline. A SPIRIT-compliant schematic diagram will be appended in the final version.*

### 3.6 Sample Size

This power calculation is based on Blanco et al., 2020, the most directly comparable study. Our trial defines the ITT population as manuscripts reaching first revision (a complete-case framework), so we base the effect size on Blanco’s complete-case results (n=18): adjusted mean difference of 1.75 items on a 0–8 scale (95% CI 0.80 to 2.75), with SDs of 1.00 (intervention) and 1.35 (control), yielding a pooled SD of ∼1.19 and a standardized effect size of d ≈ 1.5. As there are more assessed items in our study than in Blanco’s (only 8), this power analysis offers 3 scenarios: an optimistic scenario in which items are reported independently, leading to a square root of the number of items increase, a primary scenario in which they are correlated, and a conservative scenario in which there are diminishing returns on marginal items.

For simplicity, we assume the OLS model described above. Required sample sizes for 90% power, two-sided alpha = 0.05. Covariate adjustment assumes R² = 0.30 from pre-specified covariates (primarily number of improvable items). Note: R² = 0.30 is an assumption (r ≈ 0.55 between baseline and outcome), not a value extracted from Blanco et al., who did not report R². Attrition assumes 25% of randomized manuscripts do not reach first revision (consistent with Blanco et al.). All totals are rounded to a multiple of 4 for 1:1 allocation with a block size of 4.

| Scenario | d | N/arm (unadj.) | N/arm (adj.) | Total ITT (adj.) | Randomize/arm (25% attrit.) | Total to randomize |
| --- | --- | --- | --- | --- | --- | --- |
| A: Optimistic | 2.4 | 4 | 3 | 6 | 4 | 8 |
| **B: Primary** | **1.5** | **10** | **7** | **14** | **10** | **20** |
| C: Conservative | 0.9 | 26 | 19 | 38 | 26 | 52 |

To account for additional censoring from publishing the results of this study before all manuscripts have been resubmitted as first revision, we commit to a sample size of 40, which 20 randomized in each arm.

### 3.7 Recruitment

All eligible RCT submissions to participating journal(s) during the recruitment window will be considered. We anticipate 6–12 months of recruitment to reach the target sample size. The trial will close enrollment once the required number of first revisions has been accrued. If recruitment is slower than expected, additional participating journals may be invited. Recruitment progress will be monitored by the trial oversight group.

## 4. Methods: Assignment of Interventions

### 4.1 Allocation

#### Sequence generation

A computer-generated random allocation sequence will be used, with permuted block randomization (block sizes of 4) and a 1:1 allocation ratio. Details of the block sizes will be documented as an attachment to this protocol.

#### Allocation concealment mechanism

The allocation sequence will be implemented via the python script that generates the Rohe Nordberg Report and will assign manuscripts to arms at the point of eligibility confirmation, after desk screening is complete. The sequence is concealed from all personnel until the moment of assignment.

#### Implementation

The allocation sequence will be generated by Auden Nordberg Krauska. Editorial staff will confirm manuscript eligibility (passing desk screening), at which point the system automatically assigns the manuscript to an arm. A separate LLM call that confirms eligibility does not have foreknowledge of the upcoming allocation. Another separate LLM call generates the Rohe Nordberg report.

### 4.2 Blinding (Masking)

Due to the nature of the intervention, authors, editors and AEs in the intervention arm are necessarily aware of their assignment (they receive the Rohe Nordberg Report). However, the following stakeholders are blinded:

- **Peer reviewers:** Are not informed of the arm assignment.
- **Outcome assessors (human reviewers of diff files):** The diff files are stripped of any identifying information that could reveal arm assignment. Reviewers do not know whether the AE received the Rohe Nordberg Report. Files are identified only by a blinded study code. The process for generating and de-identifying the diff files will be managed by a team member who is not involved in the outcome evaluation.
- **Data analysts:** The statistician will receive a coded dataset (Arm A/Arm B) and will conduct the primary analysis blinded to which code represents the intervention.

Unblinding is not applicable in the traditional sense, as no emergency unblinding procedures are needed. Arm assignments will be unblinded to the data analyst only after the primary analysis is finalized and locked.

## 5. Methods: Data Collection, Management, and Analysis

### 5.1 Data Collection Methods

Data will be collected from the following sources:

- **Manuscripts and diff files:** The original submission and the first revision. For each manuscript, a structured document showing the textual differences between the original submission and the first revision will be generated programmatically. These files will serve as the primary input for human outcome assessment.
- **Human reviewer assessments:** Blinded human reviewers will evaluate each diff file using a standardized evaluation form. For each CONSORT item in the Methods and Results sections, reviewers will record whether reporting improved, was maintained, or reduced. Reviewers will receive a detailed coding manual and undergo calibration training using exemplar files before beginning assessments. Each file will be independently assessed by at least two reviewers.
- **LLM page-reference verification (secondary outcome):** For the secondary outcome of page-reference accuracy, the LLM-based system will verify whether authors’ stated page numbers match the location of relevant content in the revised manuscript.
- **Editorial system data:** Rejection/acceptance decisions, dates of submission and decisions, and author resubmission status are extracted from the journal’s online submission system.
- **Revision extent:** Word count of differential changes (insertions + deletions) computed programmatically by comparing the original submission to the first revision.

In this trial, the analog of participant retention is whether authors submit a first revision after receiving the editorial decision. Manuscripts for which authors never resubmit are not simply lost to follow-up — the non-resubmission rate is itself a pre-specified secondary outcome (Section 3.4), as the intervention could plausibly affect authors’ willingness to revise (e.g., a longer or more detailed revision request might discourage resubmission, or conversely, clearer guidance might encourage it). All randomized manuscripts will be tracked through ScholarOne regardless of whether a revision is received. For manuscripts that are not resubmitted, the date of the editorial decision, the decision type, and whether the AE shared the Rohe Nordberg Report (intervention arm) will be recorded. For the primary outcome analysis, only manuscripts that reach first revision are included in the ITT population; non-resubmitted manuscripts are analyzed through the non-resubmission rate and the time-to-revision survival analysis (in which they are censored).

### 5.2 Data Management

Human reviewer assessments are entered into a standardized electronic data capture form with built-in range checks, logic checks, and completeness validation. Diff files, reviewer forms, and editorial outcome data are stored in a secure, access-controlled database and linked via a unique blinded manuscript identifier. Access is limited to authorized members of the research team.

### 5.3 Statistical Methods

#### Primary analysis

The primary pre-specified analysis will use a covariate-adjusted regression model. The primary outcome — the number of CONSORT Methods and Results items per manuscript for which human reviewers judge that reporting improved at first revision — will be modeled as a function of arm assignment and a set of pre-specified baseline covariates: number of improvable items (the count of CONSORT Methods and Results items not classified as fully reported at baseline, derived from the LLM’s evaluation of the original submission applied to both arms), word count of the original manuscript, number of authors, country of corresponding author (categorized as UK, other Europe, North America, Asia, other), whether a CONSORT checklist was uploaded at original submission, and number of tables and figures. All covariates are derived from ScholarOne data at the time of original submission, before the intervention.

The model family will be finalized in a SAP amendment after completion of the pilot study but before any outcome data from the main trial are unblinded. Two candidate models are:

- OLS regression with inference based on the nonparametric bootstrap (minimum 10,000 resamples), yielding an adjusted mean difference in number of items improved with a 95% bootstrap percentile confidence interval.
- Negative binomial regression with log link, yielding an incidence rate ratio. Marginal mean differences will be computed via g-computation for interpretability, with bootstrap confidence intervals.

The model will be selected by a mechanical decision rule applied to pilot data: an intercept-only negative binomial model will be fit to the pilot outcome data (pooled across arms, without unblinding). If the estimated dispersion parameter exceeds 1.5, or if a likelihood ratio test comparing Poisson vs. negative binomial yields p < 0.10, negative binomial will be selected.

Otherwise, OLS will be selected. The unchosen model will be reported as a pre-specified sensitivity analysis. The SAP amendment documenting the model choice, with supporting pilot diagnostics, will be filed and timestamped before main trial unblinding.

Because manuscripts with fewer items fully reported at baseline have more "room to improve," the number of improvable items is expected to be the strongest prognostic covariate. This variable will be derived from the total number of items subtracted by the LLM’s evaluation of the original submission (applied to both arms). The number of improvable items will be entered into the model as log(1+improvable items) to account for non-linearity.

An unadjusted comparison (two-sample t-test or Wilcoxon rank-sum test) will also be reported for transparency but is not the basis for primary inference. The covariate-adjusted model is the single primary analysis; no multiplicity adjustment is required.

#### Secondary analyses

- Correct page references: The mean proportion of items with verified correct page numbers will be compared between arms using a Wilcoxon rank-sum test. Mean difference with bootstrap 95% confidence interval will be reported.
- Editorial decisions: Rejection rates compared by Fisher’s exact test with risk difference and 95% confidence interval; time to final decision compared by Wilcoxon rank-sum test with Hodges-Lehmann median difference and 95% confidence interval.
- Author response time: Time to resubmission compared by Wilcoxon rank-sum test with Hodges-Lehmann median difference and 95% confidence interval.
- Extent of revisions: Word count of differential changes compared between arms using a Wilcoxon rank-sum test with Hodges-Lehmann median difference and 95% confidence interval.
- AE sharing of the report (intervention arm only): The proportion of intervention-arm manuscripts for which the AE shared the Rohe Nordberg Report with authors (via the "shared with author" toggle in ScholarOne) will be reported with a 95% confidence interval. No between-arm comparison is performed; this characterizes intervention uptake.

Secondary outcomes are considered supportive and exploratory. No formal multiplicity adjustment will be applied; all secondary p-values will be reported as nominal.

#### Additional analyses

- Per-protocol sensitivity analysis: The primary model will be re-run excluding intervention-arm manuscripts where the AE did not share the Rohe Nordberg Report with authors (as determined by the ScholarOne "shared with author" toggle). This is the pre-specified per-protocol criterion because sharing the report with authors represents the minimum threshold at which the intervention can plausibly influence the primary outcome.
- Unchosen candidate model: The candidate model not selected by the pilot-data decision rule will be fit to the main trial data and reported as a sensitivity analysis.
- Poisson regression: A Poisson regression model with the same covariate set will be fit as an additional sensitivity check on the variance assumption.
- AE clustering sensitivity analysis: A mixed-effects model with AE as a random intercept will be fit to account for potential clustering of manuscripts within AEs.
- Inter-rater reliability: Cohen’s kappa will be computed overall, per CONSORT item, and separately by arm.
- Alternative reviewer disagreement handling: Sensitivity analyses using (a) the more conservative rating ("not improved" if either reviewer said "not improved") and (b) the more liberal rating ("improved" if either reviewer said "improved") in place of the consensus/adjudicated rating.
- Pre-specified subgroup analyses will explore treatment effect heterogeneity by number of improvable items (below vs. above median), country region (UK vs. non-UK), manuscript word count (below vs. above median), and whether a CONSORT checklist was submitted (yes vs. no). These will be conducted by adding interaction terms to the primary model. All subgroup analyses are exploratory; interaction p-values will be reported with a threshold of p < 0.10 to flag potential heterogeneity.
- Exploratory item-level analysis: For each individual CONSORT Methods and Results item, the proportion of manuscripts showing improvement will be compared between arms using Fisher’s exact test, displayed in a forest plot.

#### Analysis population and missing data

The primary analysis will follow the intention-to-treat (ITT) principle: all randomized manuscripts that reach first revision will be analyzed in the arm to which they were randomized, regardless of whether the AE shared or used the report. Manuscripts that do not reach first revision (e.g., author withdrawal) will be described but excluded from the primary analysis. Where two human reviewers disagree on a given item, the resolved consensus rating will be used. Missing primary outcome data is not anticipated: the primary outcome is derived from diff files comparing the original submission and first revision, both of which are stored in ScholarOne for every manuscript that reaches first revision. Because diff file generation is a deterministic programmatic operation on documents already in hand, there is no mechanism by which primary outcome data would be unavailable for a manuscript in the ITT population.

## 6. Methods: Monitoring

### 6.1 Data Monitoring

Given that this trial involves an editorial intervention with no direct risk to patients or trial participants (the unit of randomization is a manuscript, not a person), a formal independent Data Monitoring Committee (DMC) is not considered necessary. Oversight of data quality, recruitment progress, and protocol adherence will be performed by the trial oversight group.

An interim analysis may be necessary to facilitate Auden Nordberg Krauska’s dissertation and PhD graduation. An analysis of all available data on August 3, 2026 will be performed.

### 6.2 Harms

This trial does not involve direct patient contact, pharmaceutical agents, or medical devices, and therefore traditional adverse event monitoring is not applicable. Potential harms are limited to editorial process disruptions (e.g., delays caused by additional review steps, AE dissatisfaction with the tool). These will be monitored through communication with the editorial board and tracked time-to-decision metrics. Any significant negative effects on editorial workflow will be reported to the trial oversight group.

### 6.3 Auditing

Protocol adherence will be audited by the trial oversight group at quarterly intervals. Auditing will verify: (a) that randomization was correctly implemented, (b) that intervention-arm manuscripts generated the Rohe Nordberg Report, (c) that diff files were correctly generated and de-identified before reaching human reviewers, and (d) that outcome assessments were conducted according to the coding manual. This auditing process is independent from the investigators conducting the primary analysis.

## 7. Ethics and Dissemination

### 7.1 Research Ethics Approval

This trial involves an intervention on editorial processes rather than on human participants. Ethics review was sought from the University of Wisconsin-Madison IRB and a certificate of not human subjects research was granted on March 11, 2026.

### 7.2 Protocol Amendments

Any important protocol modifications (changes to eligibility criteria, outcomes, analyses, or other key design elements) will be documented as formal amendments, with a version number, date, and summary of changes. Amendments will be communicated to the relevant ethics committee/IRB, the trial registry, participating journals, and the trial oversight group. A complete version history will be maintained as an appendix to this protocol.

### 7.3 Consent or Assent

Because the unit of randomization is a manuscript and the intervention targets the editorial review process (not human participants), traditional informed consent from trial participants is not applicable. Editors and AEs will be informed about the trial and their potential role in it.

Authors will not be individually consented, as the intervention operates within the normal scope of editorial decision-making. The IRB approved the ethical justification for waiver of consent.

Not applicable. No biological specimens or participant-level clinical data are collected.

### 7.4 Confidentiality

All manuscript data will be handled in accordance with the journals’ existing confidentiality policies. Manuscripts are identified by a unique blinded study code; no personally identifiable author information will be included in the analytical dataset or in the diff files provided to human reviewers. LLM outputs, reviewer assessments, and editorial decision data are stored on secure, access-controlled servers. LLMs will be accessed through the journal’s Google Vortex with zero data retention. Only de-identified, aggregated data will be reported in publications.

Data sharing agreements with participating journals will specify confidentiality protections.

### 7.6 Access to Data

The principal investigators will have full access to the final trial dataset. No contractual agreements limit investigator access to data. Post-completion, de-identified data will be shared in an open repository, subject to editorial confidentiality protections and any restrictions imposed by data-sharing agreements with participating journals.

### 7.7 Ancillary and Post-Trial Care

Not applicable. This trial involves no direct patient contact. No ancillary care, compensation for harm, or post-trial care provisions are required. Following trial completion, participating journals may choose to adopt or discontinue the LLM-based tool at their discretion.

### 7.8 Dissemination Policy

Results will be submitted for publication in a peer-reviewed journal (e.g., The BMJ). Findings will also be presented at relevant conferences. Results will be reported in the trial registry regardless of the direction or magnitude of findings. There are no publication restrictions imposed by the sponsor or funder.

The full protocol, de-identified manuscript-level dataset, statistical code, and the coding manual used by human reviewers will be made publicly available in an open repository (Open Science Framework) following publication of the primary results, subject to editorial confidentiality protections.

## 8. Appendices

### 8.1 Informed Consent Materials

Not applicable. As detailed in Section 7.3, traditional informed consent is not required for this trial.

### 8.2 Biological Specimens

Not applicable. No biological specimens are collected in this trial.

## 9. Discussion

By providing editors and AEs with a structured Rohe Nordberg Report, incomplete reporting may be more easily identified and addressed before final acceptance. Using blinded human reviewers as the primary outcome assessors, rather than relying on automated LLM-based evaluation, ensures that the outcome measurement itself is both rigorous and free from potential systematic biases inherent in automated classification. The use of diff files allows reviewers to focus directly on what changed between the original submission and the first revision, providing a clear and targeted evaluation of improvement.

This trial has several strengths. The randomized design eliminates the concern for confounding. Blinding of outcome assessors (human reviewers) and data analysts reduces assessment bias. The focus on Methods and Results CONSORT items covers the sections most critical for replicability and evidence synthesis. The pragmatic nature of the intervention enhances external validity.

Limitations include the inability to blind editors and AEs to their arm assignment and potential variability in how they incorporate the report into their editorial feedback. Exposure to the Rohe Nordberg Report for intervention-arm manuscripts may lead editors and AEs to internalize CONSORT reporting expectations and apply heightened scrutiny to subsequent control-arm manuscripts. This contamination is expected to be conservative, and so a statistically significant finding remains valid and should be interpreted as a lower bound of the true intervention effect. Human outcome assessment introduces potential subjectivity, which is mitigated by standardized training, a detailed coding manual, independent dual review, and formal inter-rater reliability assessment. Additionally, the trial is conducted within a specific journal or set of journals, which may limit generalizability to other editorial environments.

Should the intervention arm significantly outperform control, it may encourage further adoption of LLM-based editorial aids across academic publishing, with potential benefits for the completeness of the scientific literature.

## Data Availability

Data for this study has not yet been collected, as it is a protocol.

## Abbreviations

AE: Associate Editor
CONSORT: Consolidated Standards of Reporting Trials
DMC: Data Monitoring Committee
ICMJE: International Committee of Medical Journal Editors
IRB: Institutional Review Board
ITT: Intention-to-Treat
LLM: Large Language Model
RCT: Randomized Controlled Trial
RoB 2: Risk of Bias 2
SPIRIT: Standard Protocol Items: Recommendations for Interventional Trials
WHO: World Health Organization

## Authors’ Contributions

[To be completed.]

